# Age-specific severity of SARS-CoV-2 in February 2020 – June 2021 in the Netherlands

**DOI:** 10.1101/2023.02.09.23285703

**Authors:** Pieter T. de Boer, Jan van de Kassteele, Eric R.A. Vos, Liselotte van Asten, Dave A. Dongelmans, Arianne B. van Gageldonk-Lafeber, Gerco den Hartog, Agnetha Hofhuis, Fiona van der Klis, Dylan W. de Lange, Lenny Stoeldraijer, the RIVM COVID-19 epidemiology and surveillance team, Hester E. de Melker, Eveline Geubbels, Susan van den Hof, Jacco Wallinga

## Abstract

**Background:** Severity of SARS-CoV-2 infection may vary over time. Here, we estimate age-specific risks of hospitalization, ICU admission and death given infection in the Netherlands from February 2020 to June 2021.

**Methods:** A nationwide longitudinal serology study was used to estimate numbers of infections in three epidemic periods (February 2020 – June 2020, July 2020 – February 2021, March 2021 – June 2021). We accounted for reinfections and, as vaccination started in January 2021, breakthrough infections among vaccinated persons. Severity estimates were inferred by combining numbers of infections with aligned numbers of hospitalizations and ICU admissions from a national hospital-based registry, and aligned numbers of deaths based on national excess all-cause mortality estimates.

**Results:** In each period there was a nearly consistent pattern of accelerating, almost exponential, increase in severity of infection with age. The rate of increase with age was highest for death and lowest for hospitalization. In the first period, the overall risk of hospitalization, ICU admission and death were 1.5% (95%-confidence interval [CI] 1.3-1.8%), 0.36% (95%-CI: 0.31-0.42%) and 1.2% (95%-CI: 1.0-1.4), respectively. The risk of hospitalization was higher in the following periods, while the risk of ICU admission remained stable. The risk of death decreased over time, with a substantial drop among ≥70-years-olds in February 2021 – June 2021.

**Conclusion:** The accelerating increase in severity of SARS-CoV-2 with age remained intact during the first three epidemic periods in the Netherlands. The substantial drop in risk of death among elderly in the third period coincided with the introduction of COVID-19 vaccination.

## Introduction

The severity of Severe Acute Respiratory Syndrome Coronavirus 2 (SARS-CoV-2) infection – defined as the proportion of infections resulting in hospitalization, ICU admission, and death – is an important indicator to guide pandemic response measures and hospital capacity planning. These severity estimates could vary across epidemic periods due to changes in stress on the healthcare system, availability of curative treatments and vaccines, and the emergence of new virus variants-of-concern (VOC). The overall severity depends also on the affected population, for instance, on the distribution of infections across age-groups. This emphasizes the need for severity estimates specified by age-group and over different epidemic periods.

To evaluate the severity of infection reliably at the population level, representative serological surveys are needed. Several systematic reviews present serology-based infection fatality rates (IFR) by age, geographic region and period [1, 2]. Few European studies provide estimates of the infection hospitalization rate (IHR) and/or infection intensive care unit admission rate (IICUR]), while these indicators are arguably more relevant to policy makers. Available studies estimating the IHR and IICUR are limited to periods when the wild-type virus was dominant and vaccination had not been introduced [3-6].

As many European countries, the Netherlands was hit by a first wave of COVID-19 cases in spring 2020, followed by a second wave of cases in the autumn and early winter of 2020/2021. With the emergence of the Alpha VOC In February 2021, the second wave gradually transitioned in a third wave of cases that ended in June 2021 [7]. Here, we report age-specific estimates of the IHR, IICUR and IFR of SARS-CoV-2 for the Netherlands in three epidemic periods (February 2020 – June 2020, July 2020 – February 2021, March 2021 – June 2021), approximately covering those first three waves. We base the number of infection on a nationwide longitudinal serology, and combine this with nationwide data on SARS-CoV-2-positive hospitalizations and ICU admissions from a hospital registry, and with nationwide all-cause excess deaths estimates. We explicitly account for imperfect serological testing, differences in delay times to events, reinfections, and, as SARS-CoV-2 vaccination started in the Netherlands in January 2021, for breakthrough infections.

## Methods

### SARS-CoV-2 infections

The number of SARS-CoV-2 infections by age-group (<10, 10-19, …, ≥80 years) were based on data from a nationwide, longitudinal serology study (PIENTER Corona) among participants selected from the Dutch population registry in an age- and region-specific manner (details in [8, 9]). In this study, participants in the age-range 1-92 years were repeatedly asked to provide a self-collected fingerstick blood sample and to fill out a questionnaire. Sampling rounds used for the current study were conducted in April 2020 (2,637 samples), June 2020 (6,813 samples), September 2020 (6,093 samples), February 2021 (5,981 samples) and June 2021 (5,335 samples). Serum IgG antibodies against SARS-CoV-2 Spike-S1 antigen (S1) were quantitatively measured using a validated immunoassay [10]. Seropositive individuals were classified as uninfected in the next round, unless reinfected, which was defined by a fourfold increase in antibody concentration against S1 in unvaccinated. As the roll-out of the COVID-19 vaccination program in the Netherlands started in January 2021, seropositive individuals with a self-reported positive vaccination status in the rounds of February 2021 or June 2021 were checked for a breakthrough infection, defined as the presence of a self-reported SARS-CoV-2 PCR-test confirmation; or either becoming seropositive or having a fourfold increase in antibodies concentrations against nucleocapsid (N), a target-protein absent in COVID-19 vaccines [11]. The age-specific proportions seropositive and their 95%-confidence intervals (95%-CIs) were controlled for the survey design, weighed to match the distribution of the general Dutch population regarding sex, age, ethnic background and degree of urbanization, and were adjusted for test specifics, using a specificity of 99.9% and a sensitivity of 94.3% (95%- CI: 90.6-96.7%) [9]. Self-reported positive first SARS-CoV-2 PCR-test confirmations were not considered, as this would interfere with the adjustment for test-specifics. We used the study rounds of June 2020, February 2021 and June 2021 to distinguish three epidemic periods (February 2020 – June 2020, July 2020 – February 2021, March 2021 – June 2021). We generated 1,000 Monte Carlo realizations for the age-specific proportions seroconverted by sampling from a beta-distribution, and for each sampled proportion we generated the number of infections by sampling from a Binomial distribution using the national population sizes of 1 January 2020 [12].

### SARS-CoV-2 hospitalizations and ICU admissions

Age-specific SARS-CoV-2-positive hospital admissions (<10, 10-19, …, ≥80 years) and ICU admissions (20-29, 30-39, …, ≥80 years) by date of admission were obtained from the National Intensive Care Evaluation (NICE) COVID-19 registry (more details in [13]). This registry contains information on all hospitalized persons with a positive SARS-CoV-2 test or CT-confirmed COVID-19 for nearly all hospitals and all ICUs in the Netherlands. We included admissions since 28 February 2020. A negative Binomial generalized additive model was fitted to the daily number of hospitalizations and ICU admissions using a penalized spline to account for the date effect and day-of-the-week effects. The fitted model was used to generate 1,000 Monte Carlo realizations from the predictive distribution for hospitalizations and ICU admissions. These realizations include both the uncertainty of the parameter estimates as well as residual error (resulting from a similar sampling process as with the number of infections).

### Excess all-cause death estimates

Weekly death registrations (from Thursday to Wednesday) from Statistics Netherlands [14] by age-group (<50, 50-59, 60-69, 70-79, ≥80 years) were used to estimate the number of all-cause excess deaths. We fitted a Gaussian linear model to the baseline number of deaths per week over the past 5 years, including a linear trend for the long-term effect and harmonic terms for seasonal effects [15]. Conditional on this fit, we generated 1,000 Monte Carlo realizations from the predictive distribution of the baseline mortality. If the observed number of deaths exceeded the 97.5% upper bound of the baseline fit in a certain week, we obtained the number of excess deaths by subtracting the baseline realizations from the observed number of deaths, and otherwise set the number of excess deaths to zero. To avoid the inclusion of excess deaths from a heat wave in August 2020, we did not count excess deaths in the months July and August of 2020.

### Delay times

We accounted for differences in delay time between infection and seroconversion, and between infection and a severe event by aligning all outcomes to the symptom onset date. We used the same delay times for each period. The delay time between symptom onset and seroconversion was sampled from a gamma distribution, with a median of 12 days (IQR: 9-14) [9]. The delay times from symptom onset to severe events were sampled from an empirical distribution, based on laboratory-confirmed cases included in the Dutch national COVID-19 notification database up to 1 July 2021, and for whom the symptom onset date was available. The median delays were 5 days (IQR: 2-9 days) from symptom onset to hospitalization, 7 days (IQR: 4-11 days) from symptom onset to ICU admission, and 11 days (IQR: 7-15 days) from symptom onset to death.

### Severity ratios

For each period, numbers of hospitalizations and ICU admission per day were cumulated between the aligned start- and end-dates of the epidemic periods, and excess deaths per week between the start- and end-weeks in which these aligned dates fell. The end-dates were sampled from empirical distributions of the serological sampling dates, with medians of 15 June 2020 (Interquartile Range [IQR]: 11 June 2020 – 17 June 2020), 17 February 2021 (IQR: 15 February 2021 – 19 February 2021) and 23 June 2021 (IQR: 21 June 2021 – 28 June 2021), respectively. The start-dates were fixed, using 27 February 2020 (first notified SARS-CoV-2 infection in the Netherlands) for the first period, and the day after the median sampling date of the previous period for the next periods. Point estimates of severity (IHR, IICUR and IFR) were obtained by dividing the aligned number of outcomes (reported hospitalizations, reported ICU admissions, and mean estimate of excess deaths) by the estimated number of infections. Interval estimates were obtained by dividing the 1,000 Monte Carlo realizations of the sampled cumulative number of outcomes by the sampled number of infections, taking the 2.5% and 97.5% percentiles of the Monte Carlo realizations as the 95%-CI.

## Results

### Time course of outcomes

The time course of the outcome measures together with the median sampling dates of the used serological study rounds, show that the first period captures the first wave, the second period captures the second wave with two consecutive peaks, and the third period captures the wave with the VOC Alpha (Figure 1). The sampling rounds at the end of the first period and the third period were conducted when the epidemics had subsided to low numbers of severe events, while the sampling round at the end of the second period was in between two periods of infection but at a high level of severe events. The curves of hospitalizations, ICU admissions and excess deaths differ in magnitude but follow a similar pattern across the first and second period. In the beginning of the third period, the curve of excess deaths decreases faster than the other two curves, as hospitalizations were still high but no or limited excess deaths were detected.

**Figure 1:**
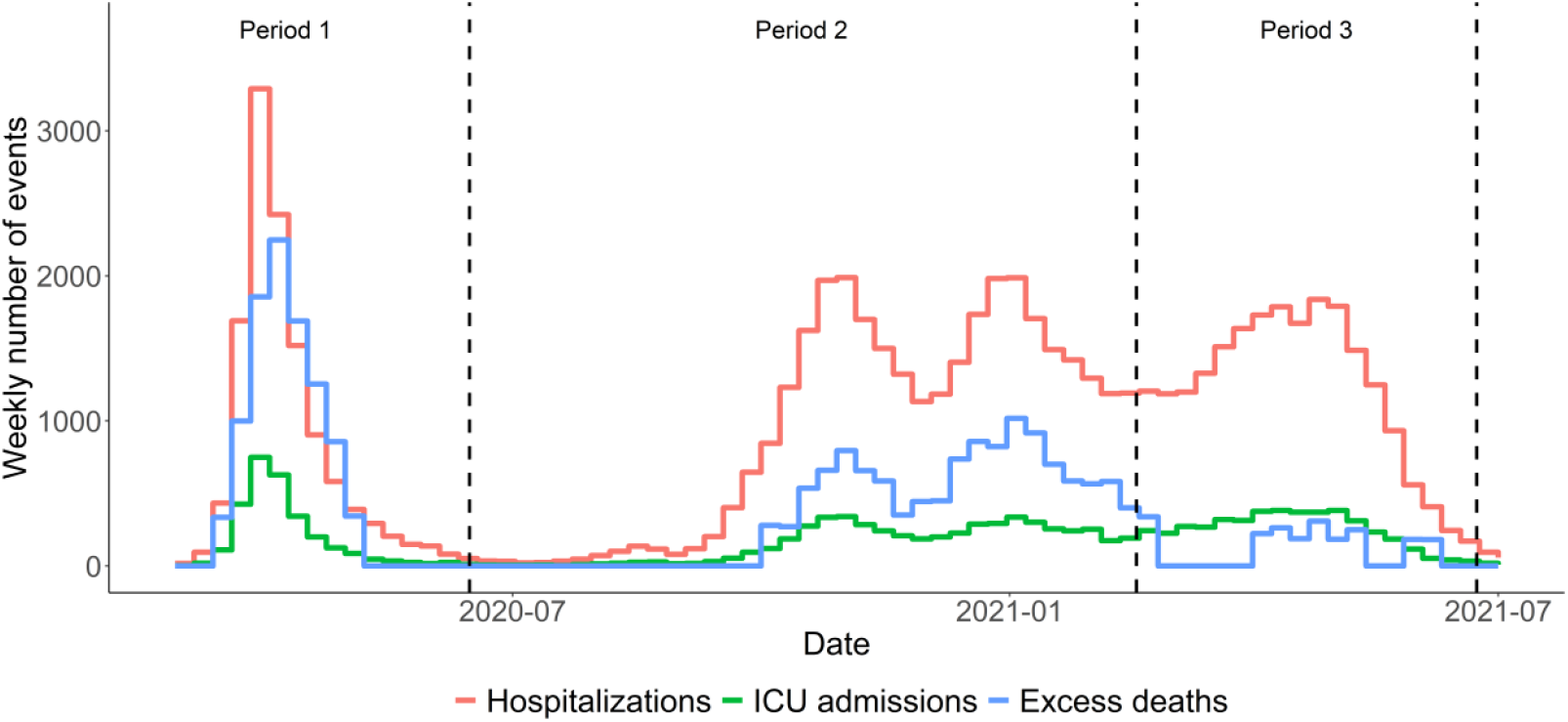
Weekly numbers of reported SARS-CoV-2-positive hospitalizations and ICU admissions by date of admission, and estimated weekly number of excess deaths in the Netherlands (mean of 1,000 simulations), from 28 February 2020 to 1 July 2021. The dashed lines represent the timing of the sampling 2^nd^, 4^th^ and 5^th^ rounds of the serological study used to distinguish the three epidemic periods.

### Time course of age-specific SARS-CoV-2 infections

Overall, the proportion of the survey population with a SARS-CoV-2 infection was 4.5% (95%-CI: 3.8-5.2%) in the first period, 8.4% (95%-CI: 7.3-9.5%) in the second period, and 7.5% (95%-CI: 6.3-8.8%) in the third period (Figure 2 and Supplemental Tables S1-S3). On the total Dutch population of 17.4 million, this corresponds to 0.79 million, 1.5 million and 1.2 million infections per period, respectively. The highest proportions of infected persons were found among 20-to 29-year-olds up to the second period, and among <20-year-olds in the third period. Only a small number of infections were measured among children aged below 10 years in the first period.

**Figure 2:**
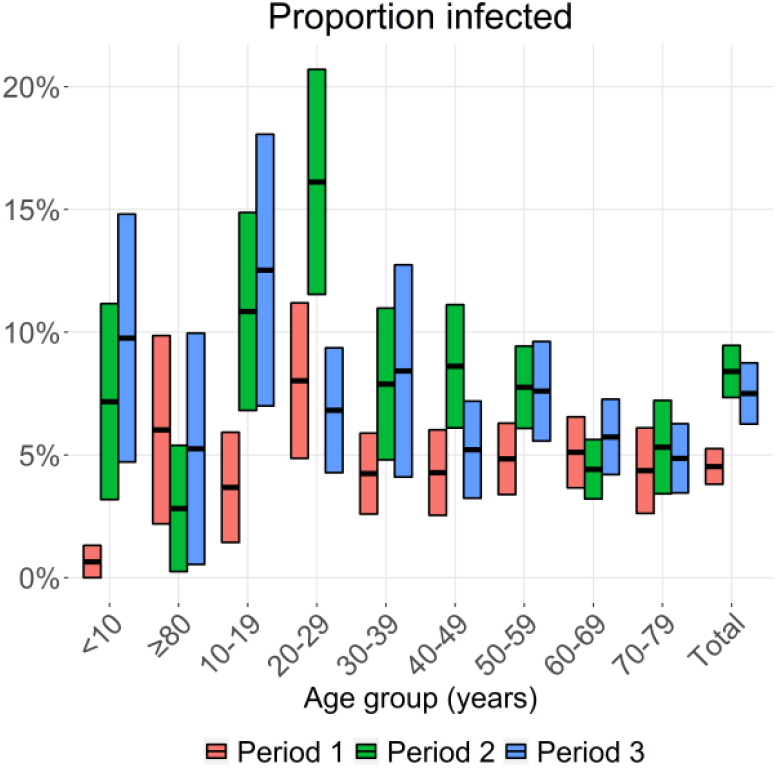
Proportion and 95% confidence interval of the study population infected per epidemic period, by age-group and overall. Period 1 is from February 2020 till June 2020, period 2 from July 2020 till February 2021, and period 3 from March 2021 till June 2021. Absolute numbers are available in Supplemental Tables S1-S3.

### Hospitalization and ICU admission per infection

The overall number of SARS-CoV-2 positive registered hospitalizations was 12,220 in the first period, 31,382 in the second period, and 22,471 in the third period. Divided by the estimated numbers of infections, these result in IHRs of 1.5% (95%-CI: 1.3-1.8%), 2.1% (95%-CI: 1.9-2.6%) and 1.7% (95%-CI: 1.5-2.0%), respectively (Figure 3A and Supplemental Tables S1-S3). Among these hospitalizations, there were 2,836 ICU admissions in the first period, 5,230 in the second period and 4,537 in the third period, resulting in IICURs of 0.36% (95%-CI: 0.31-0.42%), 0.36% (95%-CI: 0.31-0.44%) and 0.35% (95%-CI: 0.30-0.41%), respectively (Figure 3B and Supplemental Tables S1-S3). In each period, the IHR and IICUR show a nearly consistent pattern of an accelerating, almost exponential, increase with increasing age in the age-range 20-29 years to 70-79 years. The IHR was higher among 0-to 9-year-olds than 10-to 19-year-olds, and similar between 70-to 79-year-olds and ≥80-year-olds. The IICUR was lower among ≥80-year-olds compared to 70-to 79-year-olds. Over the whole study-period, the IHR doubled with every 9.0 years of increase in age, and the IICUR doubled faster, with every 7.5 years of increase in age. Compared between epidemic periods, the IHR and IICUR among 20-to 59-years were highest in the third period, although not statistically significant different from other periods in most age-groups. Above the age of 60 years, highest ratios were found during the second period.

**Figure 3:**
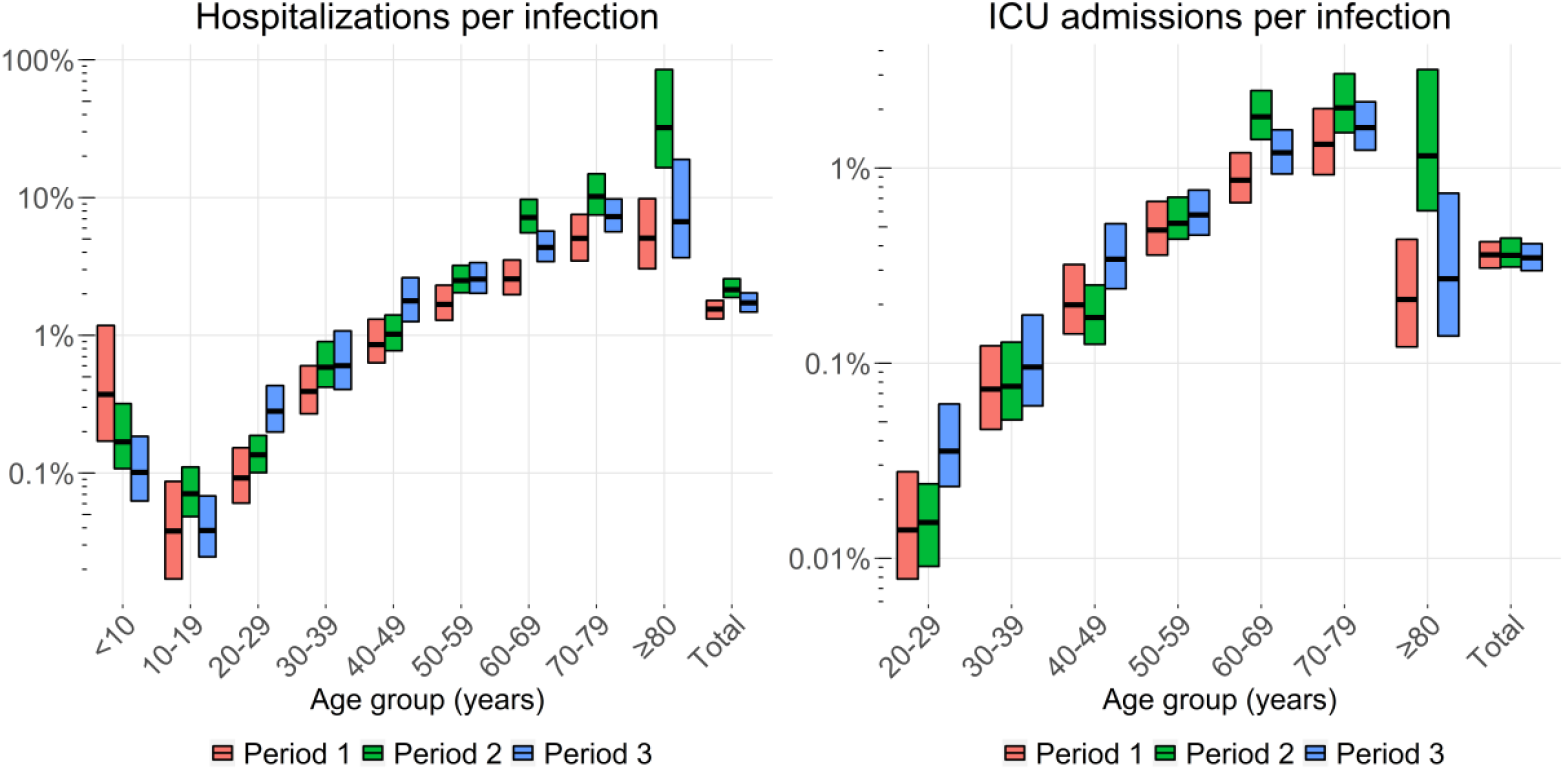
Estimated age-specific and overall infection hospitalization rates (IHR, left pane), and infection ICU admission rates (IICUR, right panel) in the Netherlands per epidemic period. Horizontal lines represent the point estimates and bars the 95% ranges using 1,000 simulations. Period 1 is from February 2020 till June 2020, period 2 from July 2020 till February 2021, and period 3 from March 2021 till June 2021. Note that the y-axes use a log-scale. Absolute estimates are available in the Supplemental Tables S1-S3.

### Excess deaths per infection

The average total number of excess deaths was estimated at 9,585 (95%-CI: 9,062-10,112) in the first period, 12,223 (95%-CI: 11,266-13,298) in the second period, and 2,512 (95%-CI: 1,897-3,147) in the third period, resulting in overall IFRs of 1.2% (95%-CI: 1.0-1.4%), 0.83% (95%-CI: 0.71-0.98%), and 0.19% (95%-CI: 0.14-0.25%), respectively. The IFR increased exponentially with age in each epidemic period (Figure 4 and Supplemental Tables S1-S3), except during the third period, where the IFR tends to drop among 60-to 69-year-olds, and significantly drops among 70-to 79-year-olds and, particularly, among ≥80-year-olds. Across the whole study period, the IFR doubled with every 4.3 years of increase of age above the age of 50.

**Figure 4:**
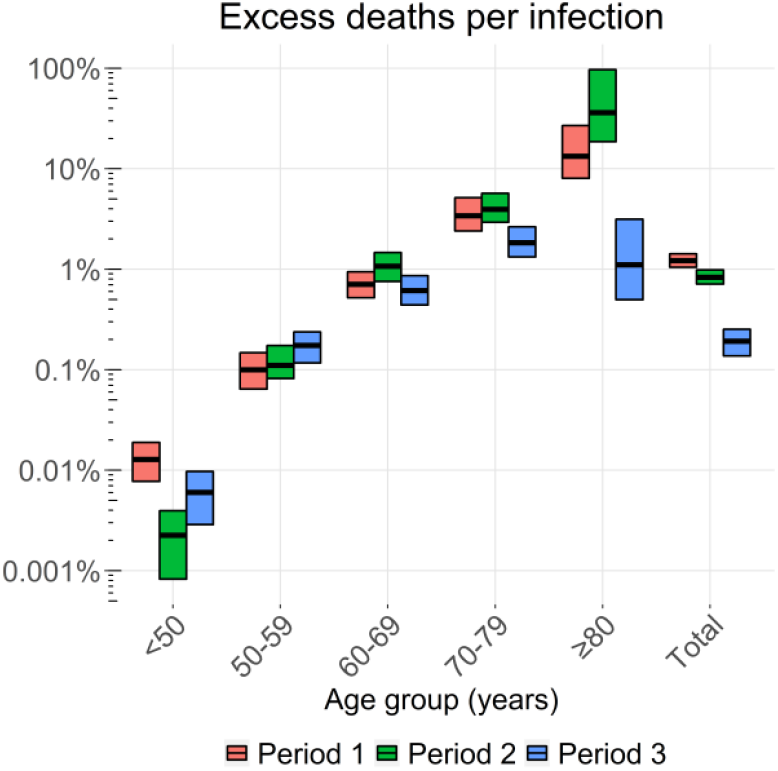
Estimated overall, and age-specific infection fatality rate (IFR) in the Netherlands per epidemic period. Horizontal lines represent the point estimates and bars the 95% ranges using 1,000 simulations. Period 1 is from February 2020 till June 2020, period 2 from July 2020 till February 2021, and period 3 from March 2021 till June 2021. Note that the y-axis uses a log-scale. Absolute numbers are available in Supplemental Tables S1-S3.

## Discussion

We analyzed the age-specific severity of SARS-CoV-2 in the Netherlands across three epidemic periods in the period February 2020 – June 2021. In each period, we found a consistent accelerating, almost exponential, increase of severity with age in the age-range 10-79 years, and this rate of increase in severity with age being determined by the severity of the outcome: the highest for death, intermediate for ICU admission, and lowest for hospitalization. The IFR dropped significantly among ≥70-years-olds in the third period.

While the IFR increased with age, the IHR was stable among ≥70-year-olds, and the IICUR was even lower among ≥80-year-olds compared to 70-to 79-year-olds. This relatively low risk of ICU admission in the eldest can be specific to the Netherlands, where frail elderly are often – after consultation with their general practitioner and family – reluctant to be admitted to the hospital and to the ICU in particular, as ICU treatment may be considered too invasive [16]. During the COVID-19 pandemic, this reluctance may have been stronger due to the high demand for hospital care.

The overall IFR decreased over time. From the first to the second epidemic period, the decrease in overall IFR was not accompanied with a decrease in age-specific IFRs, meaning that the overall decrease was mainly driven by a shift of infections to younger age-groups instead of an actual decrease in risk of death. From the second to the third period, the large drop in overall IFR was mainly driven by a significant decrease in IFR among 70-79-year-olds and, particularly, among ≥80-year-olds, which coincides with the completion of the vaccination program in this age-range. The COVID-19 vaccination program in the Netherlands started 6 weeks before the beginning of the third period, prioritizing the eldest, with approximately 80% of the ≥80-year-olds having received two doses by mid-March [17].

The decrease in overall IFR with more recent epidemic periods was not accompanied with a decrease in overall IICUR or in overall IHR. A possible explanation is that the maximum capacities in hospitals in the Netherlands were reached during the first period, and a reduction in pressure on bed capacities in following periods resulted in easing of the admission policy. From the second period onwards, treatments with dexamethasone and tocilizumab became available, as well as different forms of oxygen administration, even at home [18]. This led to a change in patient characteristics of ICU admitted patients, being on average older and having more comorbidities in the second period than in the first period [13]. In the third period, vaccination presumably had more impact on the IFR than on the IHR/IICUR, because a decent share of COVID-19-associated deaths occurred among nursing homes residents, who were prioritized for vaccination but often not admitted to the hospital.

The IHR and IICUR among 20-59-years-olds tends to be higher in the third period compared to the previous periods, although this increase is not statistically significant. This increase might be related to the emergence of the Alpha VOC, which became the dominant circulating strain in the third period. The Alpha VOC is associated with an increased severity among community-tested SARS-CoV-2 cases as compared to wild-type SARS-CoV-2 in England [19], and Denmark [20].

The estimated overall IFR of 1.2% in the Netherlands for the first period compares well with findings from other Western-European countries, estimating the IFR in the range of 0.5% to 1.8% [2, 5, 21-23]. Our finding of a decline in overall IFR from the first to the second period explained by a different age-distribution of infections was also seen in Norway [5]. The overall, and age-specific, estimates of IHR in the Netherlands tend to be lower than in other European countries, including France, Denmark and Norway [3, 5, 6], particularly among the elderly. The international comparability of IHR may be harder than IFR due to differences in hospital capacities and admission criteria. The Netherlands may have had lower IHR due to their relatively lower hospital bed capacities [24] and a more reluctant attitude towards the admission of frail elderly. This could also explain why we found the IHR to level off for above the age of 70 years, while a study from France, in which nursing home residents were excluded, reported a further increase above this age [3]. Our finding that the increase in severity with age was faster for more severe outcomes was supported by a study from France [3], and by an international meta-analysis [25].

The study has several limitations. Despite weighting the serological study sample, there could be non-participation of certain groups that have an increased risk of infection, such as ethnic minority groups [26] and nursing home residents [27], and participants of surveys may adhere better to pandemic response measures than the general population. This could also explain why we found relatively high severity estimates among ≥60-year-olds in the second period, as the estimated number of infections in this age-group was close to the number of reported cases in the national COVID-19 registry in the same period (200 thousand infections *versus* 202 thousand reported cases). This selection bias may have had less impact on estimates of infections in elderly in the first period, because the adherence to measures was generally higher [28], and in the third period, because the rolled-out vaccination programme provided protection against infection.

We used all-cause excess deaths for estimating IFR, which is an indirect measure of COVID-19 mortality that may also include deaths from indirect effects of the COVID-19 pandemic, such as delayed regular care. Also, pandemic response measures have reduced the number of deaths from other respiratory infections, such as influenza [7]. This particularly affects the excess mortality estimates of the second and third period, which overlapped with the usual respiratory illness winter season. The latter presumably also explains why no excess deaths were reported in the beginning of the third period, while substantial numbers of SARS-CoV-2 hospitalizations and ICU admissions were reported. When we compare the overall estimated numbers of excess deaths with weekly numbers of confirmed-or suspected COVID-19 deaths from cause-of-death certificates [29], we find the mean number of excess deaths to be 5% lower in the first period (9.6 thousand *versus* 10.1 thousand), 24% lower in the second period (12.2 thousand *versus* 16.0 thousand), and 54% lower in the third period (2.5 thousand *versus* 5.4 thousand). Nevertheless, the decrease in overall IFR with successive epidemic periods remained with use of cause-of-death data, resulting in estimates of 1.3% in the first period, 1.1% in the second period and 0.42% in the third period. An advantage of the use of excess death mortality is its standardization, allowing international comparisons, and its shorter delay in data availability compared to cause-of-death certificates.

Although we used a highly accurate serological assay and corrected the seroprevalence estimates for a loss in sensitivity, a small proportion of infected persons does not mount detectable antibody titers [30]. Furthermore, we could have missed reinfections or breakthrough infections from people not showing a fourfold difference in antibody concentration between successive study rounds. However, the overall seroprevalence of infection was approximately 20% in June 2021, indicating that reinfections did not substantially impact severity. Also, an unknown proportion of the reported SARS-CoV-2-positive hospitalizations were not admitted due to COVID-19, but incidentally testing positive for SARS-CoV-2 at admission, which may have led to an overestimation of COVID-19-related hospitalizations and ICU admissions, and, consequently, of severity. However, as regular care was scaled down in the Netherlands during most of the study period, we expect this would concern only a minority of admitted patients.

A major strength of our study is the use of longitudinal serology data from a large, national population-based cohort across all ages instead of relying on the (usually lower) numbers of COVID-19 notifications. Data from a high-quality national hospital-based COVID-19 registry provided us with complete data on hospitalizations and ICU admissions. We accounted for imperfect serological testing, differences in delay times to events, reinfections, and break-through infections. The findings cover a range of outcomes, a long time period and the full age range. It is consistent with previously reported severity estimates for a limited period, a limited age range, or a single outcome for other European countries. It can be used to interpolate these findings, and extrapolate them to other outcomes, periods and age groups across a range of countries.

## Conclusion

Infection-based estimates of severity of SARS-CoV-2 reveal a nearly consistent pattern of increased severity of SARS-CoV-2 with increasing age across the first three epidemic periods in the Netherlands. We confirm that the increase in severity with age was faster for more severe outcomes. Differences within age groups between periods might be explained by different circulating SARS-CoV-2 variants and the introduction of SARS-CoV-2 vaccination. We demonstrate that combining serological data from a longitudinal, nationwide survey with numbers of severe events from hospital- and death registries can provide useful insights in monitoring changes in severity of SARS-CoV-2 infection over time. Use of excess mortality for monitoring the IFR of SARS-CoV-2 appears to be challenging in periods with deviating numbers of deaths from other causes, for instance, during weeks with extreme temperatures or due to effects of pandemic response measures on other infectious diseases.

## Data Availability

All data produced in the present study are available upon reasonable request to the authors

## Statements

### Ethical statement

Ethical approval was not required for this study as all data used within this work was part of routine surveillance

## Acknowledgements

We thank the members of the RIVM COVID-19 surveillance and epidemiology team for the handling of data from various sources used in this manuscript. We gratefully acknowledge the participants of the serology study (PIENTER Corona), and the contribution of colleagues from the department of Immunology of Infectious Diseases and Vaccines (IIV) of RIVM for the laboratory analyses of the serology samples. We thank the National Intensive Care Evaluation (NICE) foundation for providing data on hospitalizations and ICU admissions, and Nicolette de Keizer of NICE for commenting on the manuscript. We thank Statistics Netherlands for providing all-cause mortality data by specified age-group.

## Conflict of interest

None

### Authors contributions

The project was conceptualised by PTdB, JvdK, ERAV, LvA, EG, SvdH and JW. Input in different parts of data collection and analysis: i) Seroprevalences: ERAV, GdH, FvdK, HEdM; ii) Hospitalizations/ICU admissions: DD, DdL, RIVM COVID-19 surveillance and epidemiology team; iii) Excess mortality: JvdK, LvA, LS; iv) Laboratory-confirmed cases: RIVM COVID-19 surveillance and epidemiology team. JvdK developed the model code, which was revised by PTdb. The initial draft of the manuscript was written by PTdb, which was critically revised by JvdK, ERAV, LvA, DD, ABvGL, GdH, AH, FvdK, DdL, LS, HEdM, EG, SvdH, JW.

### Note

Members of the RIVM COVID-19 surveillance and epidemiology team: Agnetha Hofhuis, Anne Teirlinck, Anne-Wil Valk, Carolien Verstraten, Claudia Laarman, Cheyenne van Hagen, Femke Jongenotter, Fleur Petit, Guido Willekens, Irene Veldhuijzen, Jan van de Kassteele, Janneke van Heereveld, Janneke Heijne, Kirsten Bulsink, Liselotte van Asten, Liz Jenniskens, Lieke Wielders, Loes Soetens, Maarten Mulder, Maarten Schipper, Marit de Lange, Naomi Smorenburg, Nienke Neppelenbroek, Patrick van den Berg, Priscila de Oliveira Bressane Lima, Rolina van Gaalen, Steven Nijman, Senna van Iersel, Stijn Andeweg, Susan Lanooij, Tara Smit, Thomas Dalhuisen, Don Klinkenberg, Jantien Backer, Pieter de Boer, Scott McDonald, Amber Maxwell, Annabel Niessen, Brechje de Gier, Danytza Berry, Daphne van Wees, Dimphey van Meijeren, Eric R.A. Vos, Frederika Dijkstra, Jeanet Kemmeren, Kylie Ainslie, Marit Middeldorp, Mirjam Knol, Albert Jan van Hoek, Hester de Melker, Jacco Wallinga, Rianne van Gageldonk-Lafeber, Susan Hahné, Susan van den Hof

## Supplemental materials

**Table S1:** Outcomes and severity estimates of the first epidemic period, February 2020 – June 2020.

| Age-group (years) | Population size (thousands) | Change in SARS-CoV-2 seroprevalence (%, 95%-CI) | Estimated infections (thousands, 95%-CI) | Hospital admissions | IHR (%, 95%CI) | ICU admissions | IICUR (%, 95%CI) | Estimated excess deaths (mean, 95%CI) | IFR (%, 95%CI) |
| --- | --- | --- | --- | --- | --- | --- | --- | --- | --- |
| <10 | 1,773 | 0.65 (0-1.3) | 12 (0-23) | 43 | 0.37 (0.17-1.2) |  |  |  |  |
| 10-19 | 2,002 | 3.7 (1.4-5.9) | 74 (29-119) | 28 | 0.038 (0.017-0.087) |  |  |  |  |
| 20-29 | 2,234 | 8 (4.9-11) | 179 (109-250) | 165 | 0.092 (0.06-0.15) | 25 | 0.014 (0.008-0.028) |  |  |
| 30-39 | 2,148 | 4.2 (2.6-5.9) | 91 (56-127) | 357 | 0.39 (0.27-0.6) | 67 | 0.074 (0.046-0.12) |  |  |
| 40-49 | 2,208 | 4.3 (2.5-6) | 95 (56-133) | 809 | 0.86 (0.63-1.3) | 188 | 0.2 (0.14-0.32) |  |  |
| <50 | 10,365 | 4.3 (3.4-5.3) | 446 (348-544) |  |  |  |  | 57 (36-77) | 0.013 (0.008-0.019) |
| 50-59 | 2,532 | 4.8 (3.4-6.3) | 123 (86-159) | 2,056 | 1.7 (1.3-2.3) | 590 | 0.48 (0.36-0.67) | 122 (87-159) | 0.1 (0.064-0.15) |
| 60-69 | 2,114 | 5.1 (3.7-6.6) | 108 (77-138) | 2,770 | 2.6 (2-3.5) | 938 | 0.87 (0.67-1.2) | 764 (659-869) | 0.71 (0.52-0.94) |
| 70-79 | 1,574 | 4.4 (2.6-6.1) | 69 (41-96) | 3,472 | 5.1 (3.5-7.5) | 911 | 1.3 (0.92-2) | 2,340 (2,171-2,503) | 3.4 (2.4-5.2) |
| ≥80 | 822 | 6 (2.2-9.9) | 49 (18-81) | 2,513 | 5.1 (3-9.8) | 105 | 0.21 (0.12-0.43) | 6,595 (6,110-7,071) | 13 (8-27) |
| Total | 17,408 | 4.5 (3.8-5.3) | 789 (663-914) | 12,220 | 1.5 (1.3-1.8) | 2,836 | 0.36 (0.31-0.42) | 9,585 (9,062-10,112) | 1.2 (1-1.4) |

**Table S2:** Outcomes and severity estimates of the second epidemic period, July 2020 – February 2021.

| Age-group (years) | Population size (thousands) | Change in SARS-CoV-2 seroprevalence (%; 95%-CI) | Estimated infections (thousands; 95%-CI) | Hospital admissions | IHR (%; 95%CI) | ICU admissions | IICUR (%; 95%CI) | Estimated excess deaths (mean; 95%CI) | IFR (%; 95%CI) |
| --- | --- | --- | --- | --- | --- | --- | --- | --- | --- |
| <10 | 1,773 | 7.2 (3.2-11) | 127 (56-198) | 214 | 0.17 (0.11-0.32) |  |  |  |  |
| 10-19 | 2,002 | 11 (6.8-15) | 217 (136-298) | 154 | 0.071 (0.048-0.11) |  |  |  |  |
| 20-29 | 2,234 | 16 (12-21) | 360 (258-462) | 488 | 0.14 (0.1-0.19) | 55 | 0.015 (0.009-0.024) |  |  |
| 30-39 | 2,148 | 7.9 (4.8-11) | 169 (103-236) | 994 | 0.59 (0.42-0.9) | 129 | 0.076 (0.051-0.13) |  |  |
| 40-49 | 2,208 | 8.6 (6.1-11) | 190 (135-246) | 1,941 | 1 (0.77-1.4) | 326 | 0.17 (0.13-0.25) |  |  |
| <50 | 10,365 | 8.4 (7.3-9.5) | 1,058 (886-1,229) |  |  |  |  | 24 (9-40) | 0.002 (0.001-0.004) |
| 50-59 | 2,532 | 7.8 (6.1-9.4) | 197 (154-239) | 4,909 | 2.5 (2-3.2) | 1,026 | 0.52 (0.43-0.71) | 224 (174-276) | 0.11 (0.082-0.17) |
| 60-69 | 2,114 | 4.4 (3.2-5.6) | 93 (68-119) | 6,684 | 7.2 (5.5-9.7) | 1,709 | 1.8 (1.4-2.5) | 968 (787-1,154) | 1.1 (0.76-1.5) |
| 70-79 | 1,574 | 5.3 (3.4-7.2) | 84 (54-114) | 8,531 | 10 (7.5-15) | 1,708 | 2 (1.5-3) | 3,349 (3,022-3,687) | 4 (2.9-5.7) |
| ≥80 | 822 | 2.8 (0.25-5.4) | 23 (2-44) | 7,467 | 32 (16-85) | 268 | 1.2 (0.6-3.2) | 8,374 (7,632-9,079) | 36 (19-97) |
| Total | 17,408 | 8.4 (7.3-9.5) | 1,462 (1,278-1,647) | 31,382 | 2.1 (1.9-2.6) | 5,230 | 0.36 (0.31-0.44) | 12,223 (11,266-13,298) | 0.83 (0.71-0.98) |

**Table S3:**
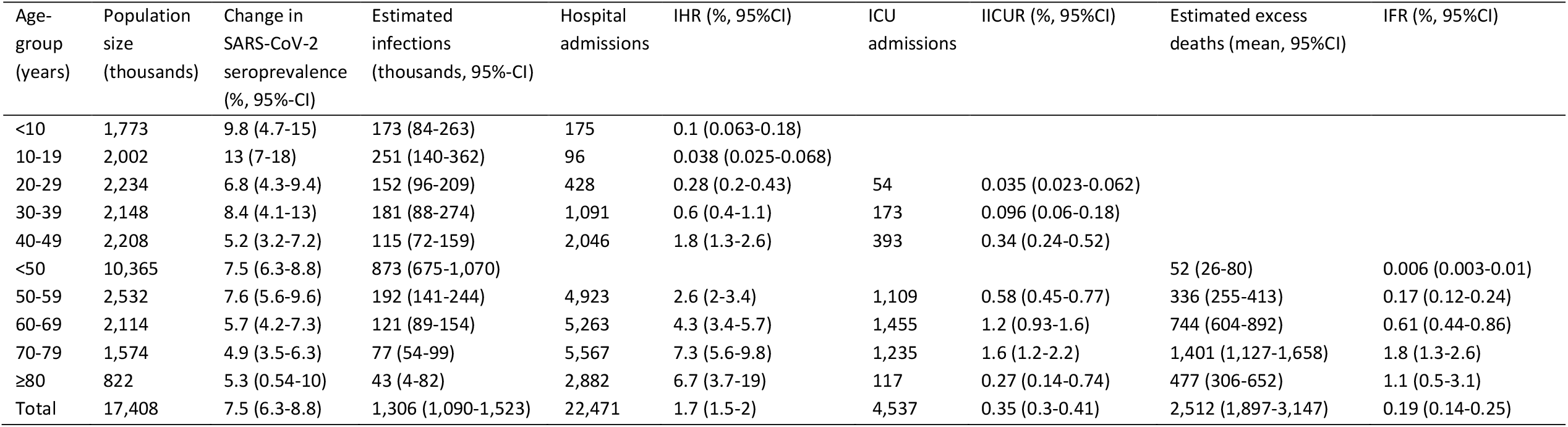
Outcomes and severity estimates of the third epidemic period, March 2021 – June 2021.

| Age-group (years) | Population size (thousands) | Change in SARS-CoV-2 seroprevalence (%; 95%-CI) | Estimated infections (thousands, 95%-CI) | Hospital admissions | IHR (%; 95%CI) | ICU admissions | IICUR (%; 95%CI) | Estimated excess deaths (mean, 95%CI) | IFR (%; 95%CI) |
| --- | --- | --- | --- | --- | --- | --- | --- | --- | --- |
| <10 | 1,773 | 9.8 (4.7-15) | 173 (84-263) | 175 | 0.1 (0.063-0.18) |  |  |  |  |
| 10-19 | 2,002 | 13 (7-18) | 251 (140-362) | 96 | 0.038 (0.025-0.068) |  |  |  |  |
| 20-29 | 2,234 | 6.8 (4.3-9.4) | 152 (96-209) | 428 | 0.28 (0.2-0.43) | 54 | 0.035 (0.023-0.062) |  |  |
| 30-39 | 2,148 | 8.4 (4.1-13) | 181 (88-274) | 1,091 | 0.6 (0.4-1.1) | 173 | 0.096 (0.06-0.18) |  |  |
| 40-49 | 2,208 | 5.2 (3.2-7.2) | 115 (72-159) | 2,046 | 1.8 (1.3-2.6) | 393 | 0.34 (0.24-0.52) |  |  |
| <50 | 10,365 | 7.5 (6.3-8.8) | 873 (675-1,070) |  |  |  |  | 52 (26-80) | 0.006 (0.003-0.01) |
| 50-59 | 2,532 | 7.6 (5.6-9.6) | 192 (141-244) | 4,923 | 2.6 (2-3.4) | 1,109 | 0.58 (0.45-0.77) | 336 (255-413) | 0.17 (0.12-0.24) |
| 60-69 | 2,114 | 5.7 (4.2-7.3) | 121 (89-154) | 5,263 | 4.3 (3.4-5.7) | 1,455 | 1.2 (0.93-1.6) | 744 (604-892) | 0.61 (0.44-0.86) |
| 70-79 | 1,574 | 4.9 (3.5-6.3) | 77 (54-99) | 5,567 | 7.3 (5.6-9.8) | 1,235 | 1.6 (1.2-2.2) | 1,401 (1,127-1,658) | 1.8 (1.3-2.6) |
| ≥80 | 822 | 5.3 (0.54-10) | 43 (4-82) | 2,882 | 6.7 (3.7-19) | 117 | 0.27 (0.14-0.74) | 477 (306-652) | 1.1 (0.5-3.1) |
| Total | 17,408 | 7.5 (6.3-8.8) | 1,306 (1,090-1,523) | 22,471 | 1.7 (1.5-2) | 4,537 | 0.35 (0.3-0.41) | 2,512 (1,897-3,147) | 0.19 (0.14-0.25) |

